# Operationalising the Centiloid Scale for [^18^F]florbetapir PET Studies on PET/MR

**DOI:** 10.1101/2022.02.11.22270590

**Authors:** William Coath, Marc Modat, M Jorge Cardoso, Pawel Markiewicz, Christopher A Lane, Thomas D Parker, Ashvini Keshavan, Sarah M Buchanan, Sarah E Keuss, Matthew J Harris, Ninon Burgos, John Dickson, Anna Barnes, David L Thomas, Daniel Beasley, Ian B Malone, Andrew Wong, Kjell Erlandsson, Benjamin A Thomas, Michael Schöll, Sebastien Ourselin, Marcus Richards, Nick C Fox, Jonathan M Schott, David M Cash, the Alzheimer’s Disease Neuroimaging Initiative

**Author notes:** Data used in preparation of this article were obtained from the Alzheimer’s Disease Neuroimaging Initiative (ADNI) database (adni.loni.usc.edu). As such, the investigators within the ADNI contributed to the design and implementation of ADNI and/or provided data but did not participate in analysis or writing of this report. A complete listing of ADNI investigators can be found at: http://adni.loni.usc.edu/wp-content/uploads/how_to_apply/ADNI_Acknowledgement_List.pdf.

## Abstract

**Purpose:** The Centiloid scale provides a systematic means of harmonising amyloid-β PET measures across different acquisition and processing methodologies. This work explores the Centiloid transformation of [^18^F]florbetapir PET data acquired on a combined PET/MR scanner and processed with methods that differ from the standard Centiloid pipeline.

**Methods:** The Standard PiB and Florbetapir Calibration datasets were processed using a standardised uptake value ratio (SUVR) pipeline with MRI parcellations from the Geodesic Information Flow (GIF) algorithm in native PET space. We generated SUVRs using whole cerebellum (GIF_WC_SUVR_) and eroded white matter (GIF_WM_SUVR_) reference regions, with and without partial volume correction (PVC). Linear regression was used to calibrate these processing pipelines to the standard Centiloid approach. We then applied the resulting transformation to 432 florbetapir scans from the Insight 46 study of mostly cognitively normal individuals aged ∼70 years, and defined Centiloid cutpoints for amyloid-β positivity using Gaussian-mixture modelling.

**Results:** GIF-based SUVR processing pipelines were suitable for conversion according to Centiloid criteria. For GIF_WC_SUVR_, cutpoints translated to 14.2 Centiloids, or 11.8 with PVC. There was a differential relationship between florbetapir uptake in WM and WC regions in Florbetapir Calibration and Insight 46 datasets, causing implausibly low Centiloid values for GIF_WM_SUVR_. Linear adjustment to account for this difference resulted in Centiloid cutpoints of 18.1 for GIF_WM_SUVR_ (17.0 with PVC).

**Conclusion:** Our results show florbetapir SUVRs acquired on PET/MR scanners can be reliably converted to Centiloids. Acquisition or biological factors can have large effects on Centiloid values from different datasets, we propose a correction to account for these effects.

## INTRODUCTION

Cerebral amyloid-β (Aβ) plaque pathology is a hallmark of Alzheimer’s disease (AD) that accumulates decades before symptom onset and can be quantified *in vivo* with PET using one of several radiotracers that selectively bind to neuritic Aβ plaques [1]. The estimation of Aβ burden using PET is crucial for accurate clinical diagnosis, the study of disease progression and for assessment of eligibility and efficacy in therapeutic trials [2]. The standardised uptake value ratio (SUVR) is a widely used semi-quantitative method for analysing and normalising Aβ-PET images acquired near steady state [3]. However, measures of SUVR are dependent on many factors, including the choice of radiotracer, target and reference regions, analysis methodology, and the data acquisition method used [4–6]. Deviations in methodology lead to results that are not easily comparable across studies or between centres.

The Centiloid Project aimed to address the issue of comparability by proposing a common scale where results of Aβ PET analyses can be harmonised using a post-hoc linear transformation [7]. Anchor points at 0 and 100 Centiloid units (CL) correspond to average SUVRs in groups of young healthy controls and patients with typical AD, respectively. The project provides guidelines on how to construct transforms from new “non-standard” approaches to the Centiloid scale, and has been used to calibrate [^18^F]-labelled Aβ radiotracers to the Centiloid scale, with the paired [^18^F]-radiotracer and reference [^11^C]Pittsburgh Compound B (PiB) images used in the calibration being made publicly available to the research community [8–11].

In this study, we explore the implementation of the Centiloid scale for data acquired on a combined PET/MR system using [^18^F]florbetapir. PET/MR scanners have only recently become widely available but provide tangible benefit in the ability to simultaneously acquire both PET and MR modalities, thus reducing the overall burden to the participant. This is particularly true for clinical research studies, where advanced imaging protocols tend to result in longer acquisition times. However, there are substantial differences between these systems and conventional PET/CT that could affect transformation to the Centiloid scale. For example, alternative methods for attenuation correction have been developed that do not require CT (Burgos et al., 2014). Other technical differences, such as longer axial field of view (FoV), may also introduce variation to SUVR measurements.

The Insight 46 cohort is a large single site sample of mostly cognitively normal individuals aged ∼70 with florbetapir PET and MRI data acquired on a combined scanner. To our knowledge the Centiloid scale transformation has not yet been applied to data collected on a combined PET/MR scanner.

## METHODS

We validated our image analysis pipeline using ‘Standard PiB’ and ‘Florbetapir Calibration’ datasets. We then assessed the suitability of the Centiloid approach for standardisation of SUVRs in an independent florbetapir dataset from Insight 46.

### Participants

Centiloid Project ‘Standard PiB’ and ‘Florbetapir Calibration’ datasets were downloaded from the GAAIN website (http://www.gaain.org/centiloid-project). The Standard PiB dataset is described in detail in Klunk et al. [7]. Briefly, the YC-0 group consists of 34 young controls and the AD-100 group is made up of 45 individuals with a diagnosis of AD. These groups form the anchor points at 0 and 100 Centiloids.

The Florbetapir Calibration dataset is described in Navitsky et al. [9] and is made up of 46 participants with clinical sub-groups of young cognitively normal (YCN; N = 13), older cognitively normal (OCN; N = 6), at-risk (N = 3), mild cognitive impairment (MCI; N = 7), possible AD (N = 3) and AD (N = 14); these subgroups are collapsed into two groups, YCN (N = 13, aged ≤ 35 years) and mixed diagnosis elder subjects (ES; N = 33, aged > 50 years).

Insight 46 is the neuroimaging sub-study of the MRC National Survey of Health and Development (NSHD), also known as the British 1946 birth cohort which initially comprised 5362 individuals born in mainland Britain during the same week in March 1946. The study protocol is described in Lane et al. [12]. In brief, 502 participants were recruited from the wider cohort and 471 underwent PET/MR scanning in London, UK. Following quality control, 432 had both MR and list-mode PET data required for this investigation (supplementary materials, section 1). Table 1 shows demographic information on each of the participant cohorts described above.

**Table 1.** Demographic characteristics in each cohort. Data regarding sex and MMSE for the Standard PiB dataset was not published in Klunk et al. [7]. APOE-e4 carrier is defined as carrying at least one apolipoprotein E e4 allele and is unknown for the following numbers in each cohort: YC-0 = 2, AD-100 = 1, YCN = 3, ES = 11, Insight 46 = 2. MMSE was unknown for 3 participants in the YCN cohort. Abbreviations: YC-0 = Young control; AD-100 = Alzheimer’s disease; YCN = Young cognitively normal; ES = Elder subjects; MMSE = Mini-mental state examination; PiB = Pittsburgh Compound B.

|  | Standard PiB |  | Florbetapir Calibration |  | Insight 46 |
| --- | --- | --- | --- | --- | --- |
|  | YC-0 | AD-100 | YCN | ES |  |
| **N** | 34 | 45 | 13 | 33 | 432 |
| **Female sex %** | - | - | 53 | 36 | 48 |
| **APOE ε4: carrier/total (%)** | 8/32 (25) | 28/44 (64) | 1/10 (10) | 14/22 (63) | 129/430 (30) |
| **Mean MMSE (SD)** | - | - | 29.5 (0.5) | 24.7 (5.1) | 29.2 (1.0) |
| **Mean Age (SD)** | 31.5 (6.3) | 67.5 (10.5) | 27.0 (4.3) | 70.2 (9.6) | 70.6 (0.7) |

### Image Acquisition

All three datasets contain static Aβ-PET images and volumetric T1-weighted MRI; full acquisition details for all datasets have been described previously (Standard PiB: [7]; Florbetapir Calibration: [9]; Insight 46: [12]). The Standard PiB dataset consists of PiB data acquired 50-70 minutes post injection on Siemens ECAT Exact HR, Siemens ECAT Exact HR+, Philips Allegro and Siemens BioGraph TruePoint TrueV scanners. The Florbetapir Calibration dataset contains both PiB (50-70 minutes) and florbetapir (50-60 minutes), all acquired in 5-minute frames on Siemens HR+, Philips Gemini TF 64 and GE Advance scanners. Data from the Insight 46 cohort were acquired on a single 3T Siemens Biograph mMR PET-MR scanner. The MR sequences included a volumetric T1-weighted MPRAGE (repetition time (TR) = 2000 ms, inversion time (TI) = 870 ms, FOV = 282 × 282 mm, 1.1 mm isotropic resolution) and a 3D T2-weighted Turbo Spin Echo (TR = 3200 ms, TE = 409 ms, FOV = 282 × 282 mm, 1.1 mm isotropic resolution). Structural MRI were visually quality checked by experienced raters. Dynamic PET data were acquired in list-mode format after intravenous injection of 370 MBq florbetapir. Static PET images from 50-60 minutes post-injection were reconstructed from list mode data on Siemens e7 tools with a 3D ordered-subset expectation-maximisation algorithm consisting of three iterations and 21 subsets, smoothed with a Gaussian kernel with 4 mm full-width at half-maximum. For attenuation correction, pCT images were created by matching morphology from an individual’s MR scans to a database of paired MR and CT scans. pCT produces results most consistent with CT compared to other methods of attenuation correction for PET/MR [13]. PET images were also reconstructed on the scanner console using ultrashort echo-time (UTE) attenuation correction which produced results that were highly correlated with pCT; see section 2 of supplementary materials.

### Imaging Analysis

To construct the Centiloid scale, Klunk et al. [7] used PiB data processed with a standard SUVR pipeline. Where a ‘non-standard’ approach is used, e.g. a different radiotracer or analysis method, values must first be calibrated to the standard approach before scaling to CL. In this work, we investigate the use of several non-standard processing methods which use (i) a different approach for structural image segmentation/parcellation; (ii) native rather than standard space analysis; (iii) a different reference region for SUVR calculation compared to the standard pipeline and; (iv) the use of partial volume correction (PVC). We compared our non-standard processing methods to the Standard Centiloid processing using the Standard PiB dataset. We then used the Florbetapir Calibration dataset to calibrate non-standard florbetapir SUVRs to PiB SUVRs processed using the standard pipeline.

We have adapted the nomenclature set out by the Centiloid project to label results from each methodology, in the following format:

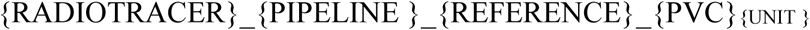

{RADIOTRACER}_{PIPELINE}_{REFERENCE}_{PVC}_{UNIT }_ Where {RADIOTRACER} is either PiB or florbetapir (FBP); {PIPELINE} is Standard Centiloid (STD) or Geodesic Information Flows (GIF); {REFERENCE} is whole cerebellum (WC) or eroded white matter (WM) region; {PVC} indicates that PVC is applied; and {UNIT} is SUVR or CL.

#### Standard Centiloid Pipeline

We implemented the Standard Centiloid processing method, denoted here as STD_WC_SUVR_, as described in Klunk et al. [7]. In brief, the individual PET frames were realigned and averaged. T1-weighted images were rigidly co-registered to the MNI152 template and the PET image was co-registered to the T1-weighted image. The T1-weighted image was then warped to MNI152 space using unified segmentation and spatial normalisation in SPM8 (revision number 4290) and the PET image was spatially normalised using the same transformation. The cortical volume of interest and whole cerebellum (WC) reference region defined on the MNI152 template were downloaded from the GAAIN website and used to calculate SUVR.

#### Non-standard GIF pipeline

We processed all three datasets described above with the following pipeline. For each individual, T1-weighted images were parcellated with Geodesic Information Flows (GIF), an automated multi-atlas propagation algorithm [14]. The T1-weighted and PET images were then co-registered using an affine block matching registration algorithm in NiftyReg [15].

The resulting transformations were used to resample the GIF parcellations into PET space. The choice of reference region, and how it is defined, has a large effect on SUVR, and there is little consensus on the optimal choice of reference region [16,17]. The whole cerebellum (WC) is a commonly adopted reference region as it is relatively spared of Aβ pathology until late stage AD [18]. Eroded white matter (WM) has also been used as a reference region as it is situated further from the edge of the axial FOV and therefore less susceptible to noise, especially in PET scanners with small axial FOV. WM as a reference has been found to increase power to detect changes in Aβ longitudinally compared to the cerebellum [19–22]. Here we calculated SUVR with two reference regions: whole cerebellum (GIF_WC_SUVR_) and subcortical white matter with an erosion of one PET voxel (GIF_WM_SUVR_).

The relatively low spatial resolution of PET means that signal from one region “bleeds” into adjacent regions, i.e. the partial volume effect; PVC attempts to minimise this [23]. PVC has been found to improve accuracy of Aβ quantification and separation of Aβ positive and negative individuals but can also introduce bias if not applied appropriately [24]. We employed the Iterative Yang method of PVC using parameters optimised for our PET/MR dataset with a Gaussian kernel of 6.8 mm FWHM and 10 iterations [23,25].

Mean SUVR values were then extracted from a large cortical composite GIF target region corresponding to the composite FreeSurfer region used by Landau et al. [6]. The composite includes frontal, cingulate, parietal, and temporal cortical regions. In total, four variants of the GIF SUVR pipeline were evaluated for calibration to the Centiloid scale: GIF_WC_SUVR_, GIF_WC_PVC_SUVR_, GIF_WM_SUVR_ and GIF_WM_PVC_SUVR_.

### Statistical Analysis

The first step of the analysis was to ensure that our in-house GIF pipeline could be calibrated to the Centiloid scale using the procedure laid out in Klunk et al. [7] for “Level 2” analysis of a non-standard method. The association between our non-standard GIF pipelines (y) and PiB_STD_WC_SUVR_ (x) in the Standard PiB dataset was assessed using linear regression, checking that the reliability threshold specified by the Centiloid project (R^2^ > 0.7) was satisfied [7]. We then calculated conversion equations for our non-standard methodology using the paired Florbetapir Calibration dataset, calibrating for differences in both radiotracer (florbetapir to PiB) as well as processing method (GIF to STD) in a single step. Florbetapir SUVRs from each GIF pipeline (y) were regressed against PiB_STD_WC_SUVR_ (x) and the reliability of the conversion process for each of the pipelines was assessed (R^2^ > 0.7). The slope and intercept of these relationships was used to transform each set of non-standard SUVRs to ^calc^PiB_STD_WC_SUVR_ (estimated values are denoted using ^calc^x_SUVR_). ^calc^PiB_STD_WC_SUVR_ values were scaled to Centiloids using equation 1.3b in Klunk et al. [7], substituting in group mean values for YC-0 = 1.00 and AD-100 = 2.07 (PiB_STD_WC_SUVR_ anchor points published in Navitsky et al. [9]). Finally, to derive direct conversion equations, Centiloid values were regressed against original SUVR values in a manner similar to Navitsky et al. [9]. Figure 1A gives a schematic overview of the Level 2 conversion process. After conversion of FBP_GIF_WM_SUVR_ to Centiloids, it became clear an additional adjustment was required, which is detailed in Figure 1B and described in the results section.

**Figure 1.**
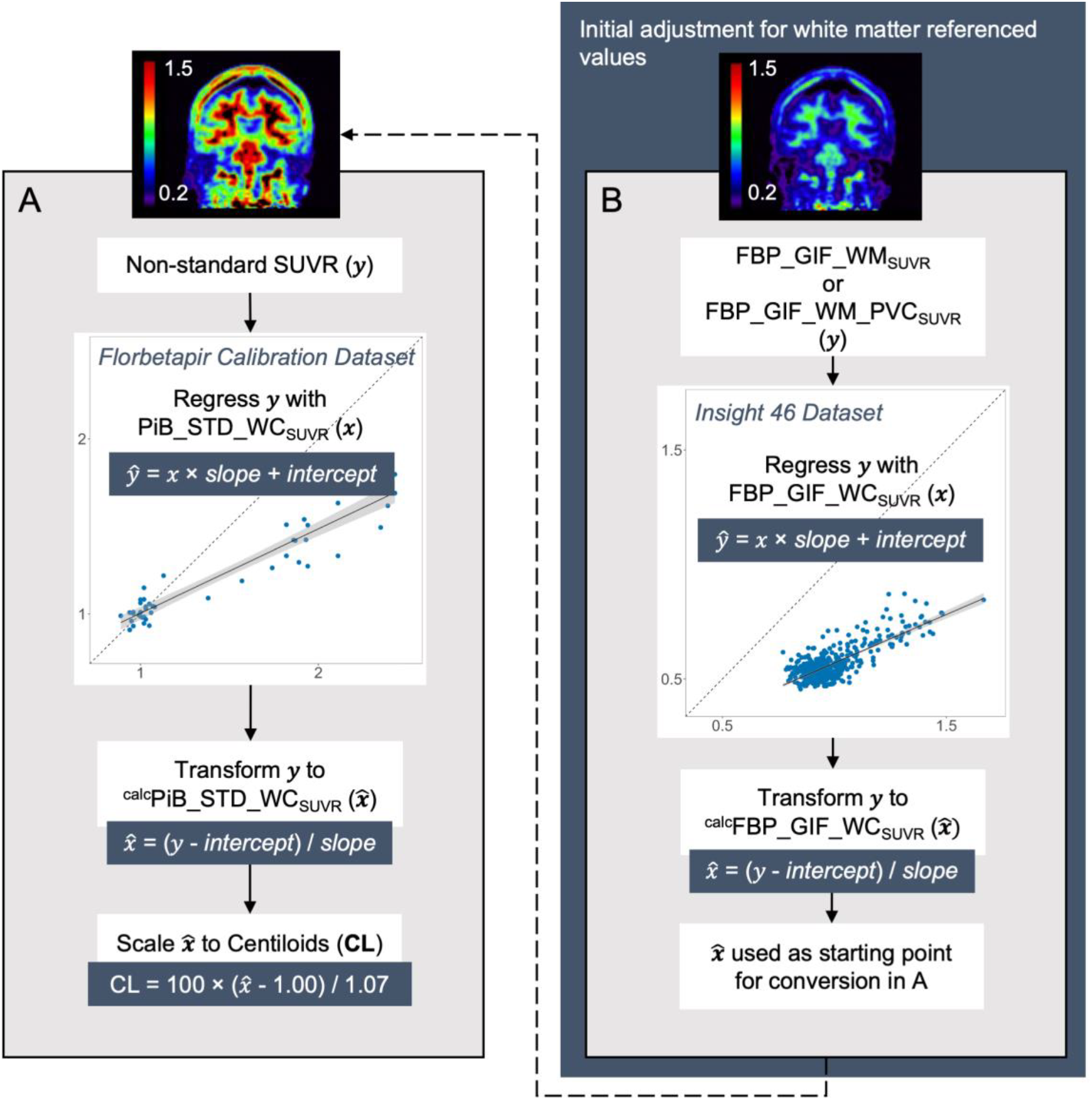
Panel A shows the process for Level 2 calibration of our pipeline methods to Centiloids (CL). Panel B shows the additional adjustment required for conversion of SUVRs using a white matter reference region.

For further comparison of processing methods, the Insight 46 dataset was also processed with a local implementation of the STD_WC_SUVR_ pipeline, which was validated through replication of the “Level 1” analysis using the Standard PiB dataset (R^2^ = 0.9994; see supplementary materials, section 3).

This conversion process was followed by assessment of the relative variance of Centiloid values generated from the non-standard pipelines. The relative variance is defined as the ratio between the SD of a non-standard approach (e.g. FBP_GIF_WC_CL_) and the SD of PiB_STD_WC_CL_ and reflects both the dynamic range and the precision of the original SUVRs, relative to the standard approach [7]. Relative variance was assessed in the YCN group of the Florbetapir Calibration dataset (N = 13). This group is assumed to have no meaningful Aβ accumulation, and thus variability should relate mostly to measurement noise and non-specific binding. We also present information on the variability of SUVRs from each approach, expressed as the coefficient of variation (CoV = 100 × SD/mean) in the YCN group.

After establishing the transformation from florbetapir SUVR to Centiloid units for each of our pipelines, we applied these transformations to florbetapir data acquired in the Insight 46 cohort. SUVR Aβ positivity cutpoints were estimated and transformed to Centiloids for each method. Cutpoints were obtained from Gaussian-mixture modelling (GMM) in the Insight 46 PET/MR dataset (MATLAB R2018a Statistics and Machine Learning toolbox). Models with one, two and three Gaussians were compared, and the two Gaussian model was selected as the optimal model based on Bayesian Information Criterion. The cutpoint value was defined as the 99^th^ percentile of the lower (Aβ negative) distribution. All other statistical analyses were performed in R version 3.6.3.

#### Supplementary analysis in ADNI dataset

The Florbetapir Calibration dataset differs from the Insight 46 dataset in both image acquisition and sample characteristics. Since the conversion equations are calculated in the Florbetapir Calibration dataset and applied to Insight 46, it is difficult to attribute any conversion issues to biological or technical aspects. To examine the Centiloid conversions in an independent age and disease stage matched PET/CT dataset, florbetapir PET images and T1-weighted MRI scans from 93 controls aged 68-72 years were downloaded from the Alzheimer’s Disease Neuroimaging Initiative (ADNI, adni.loni.usc.edu) and processed with GIF pipelines. The conversion equations were then applied to SUVRs in ADNI and Centiloid results compared (see supplementary materials, section 4).

## RESULTS

### Reliability of non-standard approaches

#### Standard PiB dataset

When comparing the non-standard GIF pipelines to the STD_WC_SUVR_ pipeline in the Standard PiB dataset, we obtained very high correlations (R^2^ between 0.91 and 0.99, see Figure 2) that were well above the established Centiloid criteria of R^2^ > 0.7. Information on the CoV and effect size of each method is provided in Table 1 in section 3 of the supplementary materials.

**Figure 2.**
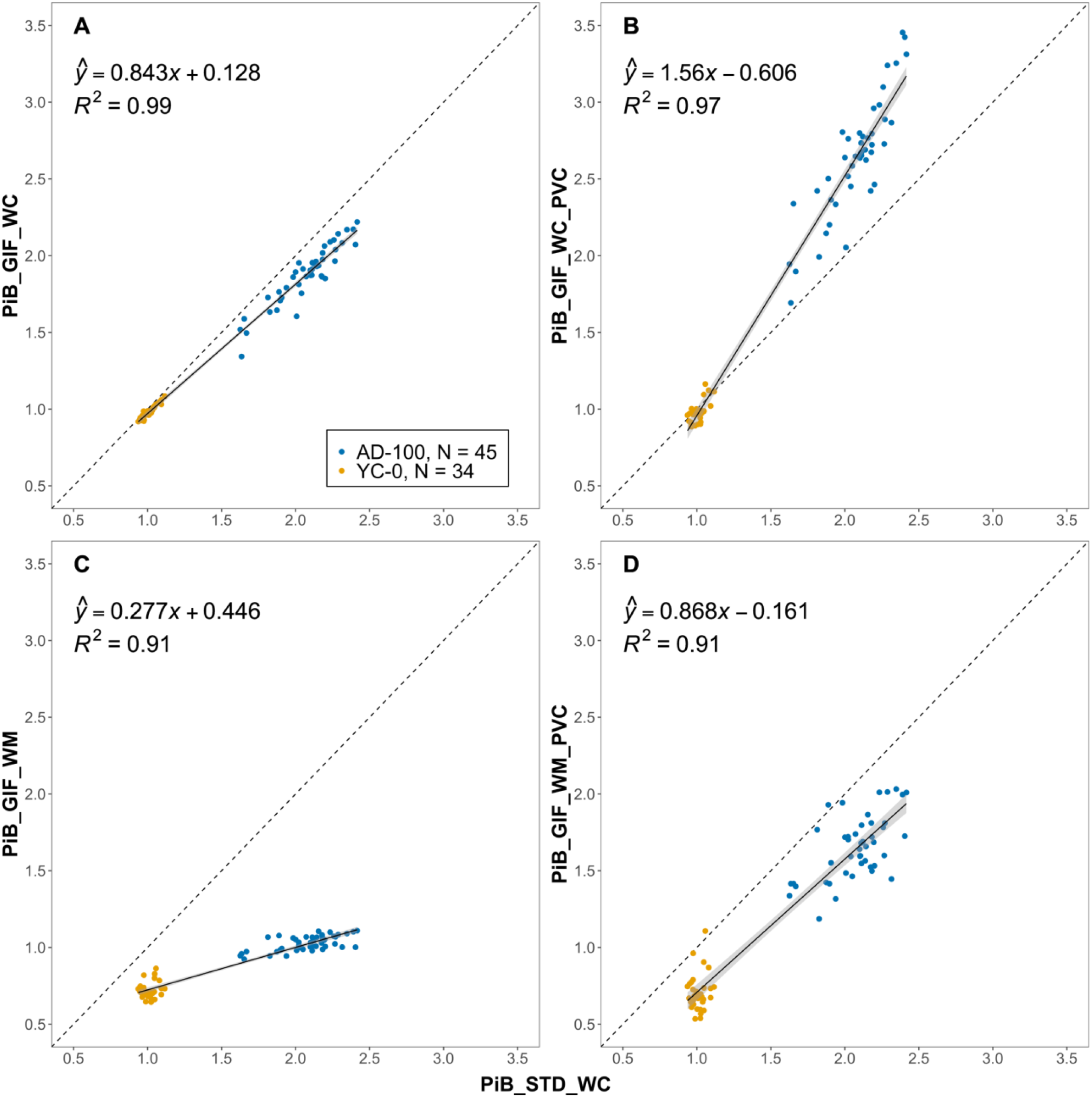
Relationship between non-standard GIF SUVR pipelines and PiB_STD_WC_SUVR_ in the Standard PiB dataset. The dashed line represents x=y and the black line is the linear regression with grey area representing 95% confidence interval. The axes were fixed in all panels to illustrate the differences in dynamic range between methods. Abbreviations: SUVR = Standardised uptake value ratio; PiB = Pittsburgh Compound B; STD_WC = standard Centiloid pipeline; GIF = geodesic information flows pipeline; WC = whole cerebellum reference; WM = eroded white matter reference; PVC = partial volume corrected; AD-100 = Alzheimer’s disease; YC-0 = Young control.

#### Florbetapir Calibration dataset

All non-standard approaches using florbetapir data reached the pre-specified Centiloid crtieria for reliability (all R^2^ > 0.7, Figure 3). Equation 2 in each panel of Figure 3 was used to convert SUVRs from each approach to ^calc^PiB_STD_WC_SUVR_ 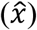 values, which were then scaled to Centiloids.

**Figure 3.**
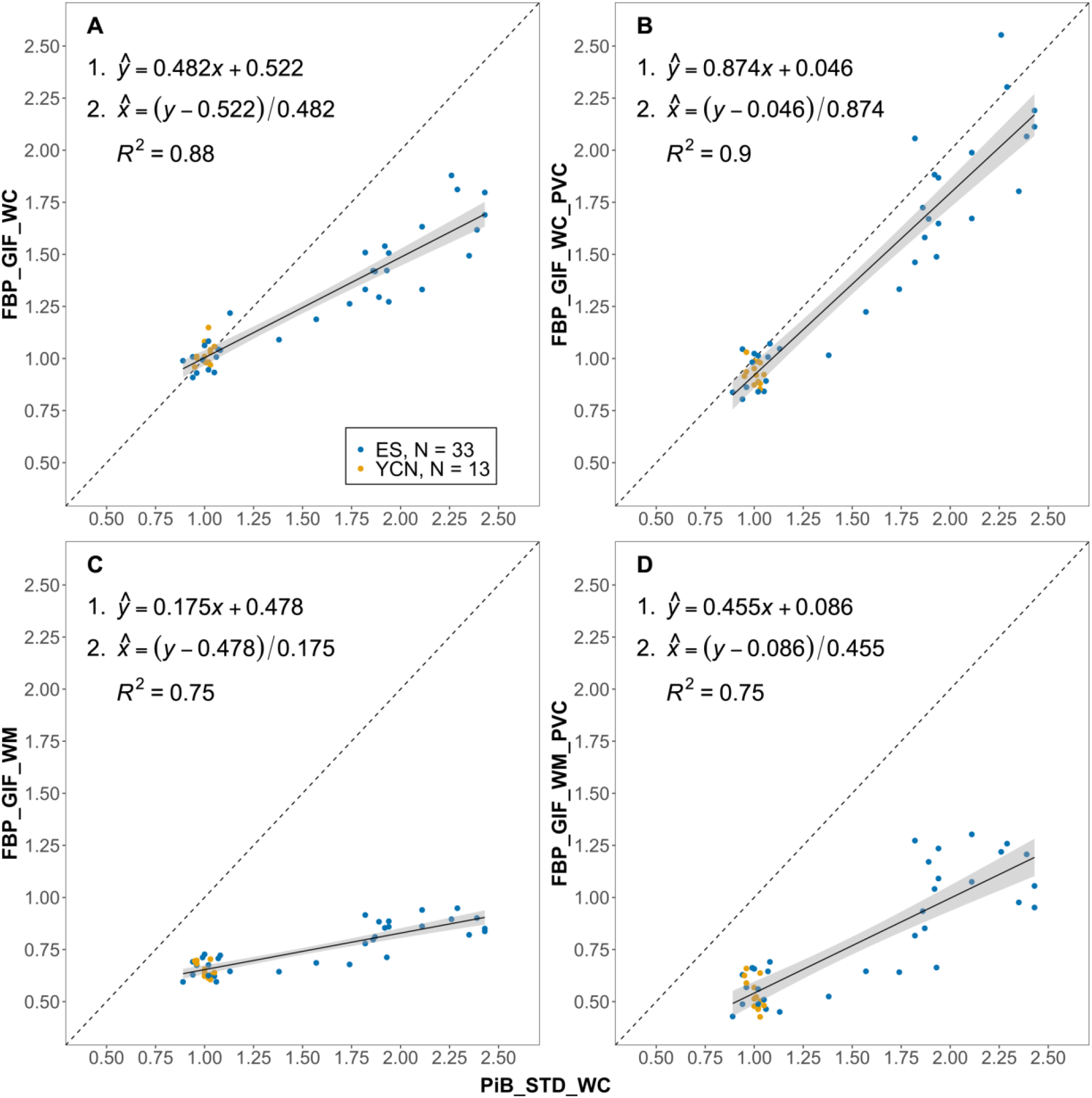
Paired florbetapir (FBP) and PiB SUVR data from the Florbetapir Calibration dataset. Plots show the relationship between FBP SUVRs (y-axis) processed using GIF_WC (A), GIF_WC_PVC (B), GIF_WM (C) and GIF_WM_PVC (D) pipelines and PiB SUVRs with STD_WC processing (all x-axis). The dashed line represents x=y and the black line is the linear regression with grey area representing 95% confidence interval. Conversion equations and R^2^ are displayed on the plots. All non-standard methods exceed the R^2^ > 0.7 reliability threshold set by Klunk et al. [7] and are therefore suitable for Centiloid conversion. Abbreviations: SUVR = Standardised uptake value ratio; FBP = florbetapir; PiB = Pittsburgh Compound B; STD = Standard Centiloid pipeline; GIF = Geodesic information flows pipeline; WC = Whole cerebellum reference; WM = Eroded white matter reference; PVC = Partial volume corrected; ES = Elder subjects; YCN = Young cognitively normal.

### Relative variance of non-standard approaches

Information on the variability of SUVRs (CoV) and Centiloids (relative variance) in YCN particpants are presented in Table 2. Reference region and PVC had an effect on the CoV in PiB data, which was lowest for PiB_STD_WC_SUVR_ and PiB_GIF_WC_SUVR_ (both 3.1%) with increasing variability with the use of a WM reference region and application of PVC. When using florbetapir data, CoV was similar for all methods (5.2 to 5.7%) apart from FBP_GIF_WM_PVC_SUVR_ which had increased variability (13.9%). The relative variance of Centiloids, defined as the SD of Centiloids from a non-standard method relative to that of PiB_STD_WC_CL_, was generally lower for PiB compared to florbetapir. WM reference region increased relative variance, and application of PVC tended to decrease it - apart from PiB_GIF_WC_PVC_CL_ where PVC caused an increase. For PiB, PiB_GIF_WC_CL_ had the lowest relative variance (0.9), whereas PiB_GIF_WM_CL_ was highest (4.5). For florbetapir data, FBP_GIF_WC_PVC_CL_ had the lowest relative variance (1.8) and it was highest in FBP_GIF_WM_CL_ (5.9).

**Table 2.**
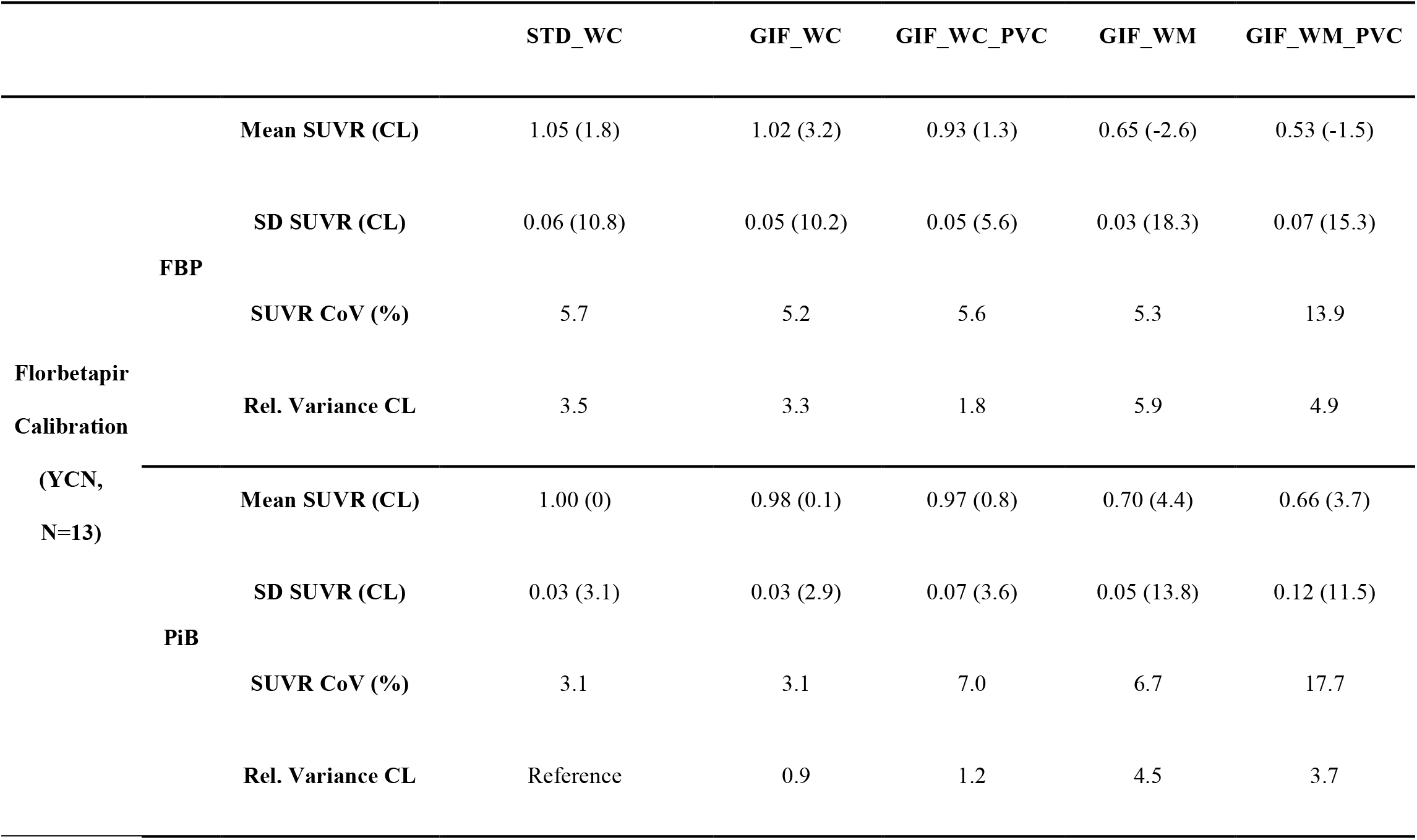
The relative variability of all processing pipelines. PiB and florbetapir SUVRs from the YCN group of the Florbetapir Calibration dataset with the CoV for each approach and relative variance of Centiloid values from each non-standard approach relative to the Standard PiB approach. Abbreviations: STD_WC = standard Centiloid pipeline; GIF = geodesic information flows pipeline; WC = whole cerebellum reference; WM = eroded white matter reference; PVC = partial volume corrected; YCN = Young cognitively normal group; FBP = Florbetapir; PiB = Pittsburgh Compound B; SD = standard deviation; SUVR = standard uptake value ratio; CL = Centiloid; CoV = Coefficient of Variance.

### Centiloid conversion of PET/MR data

#### Whole cerebellum reference region

In Insight 46 (N = 432), the Aβ positivity rates were 15.7%, 16.2% and 23.8% for STD_WC_SUVR_, GIF_WC_SUVR_ and GIF_WC_PVC_SUVR_, respectively. Direct transformation equations, cutpoint conversions and resulting distribution of Insight 46 SUVRs and Centiloids are presented in Figure 4.

**Figure 4.**
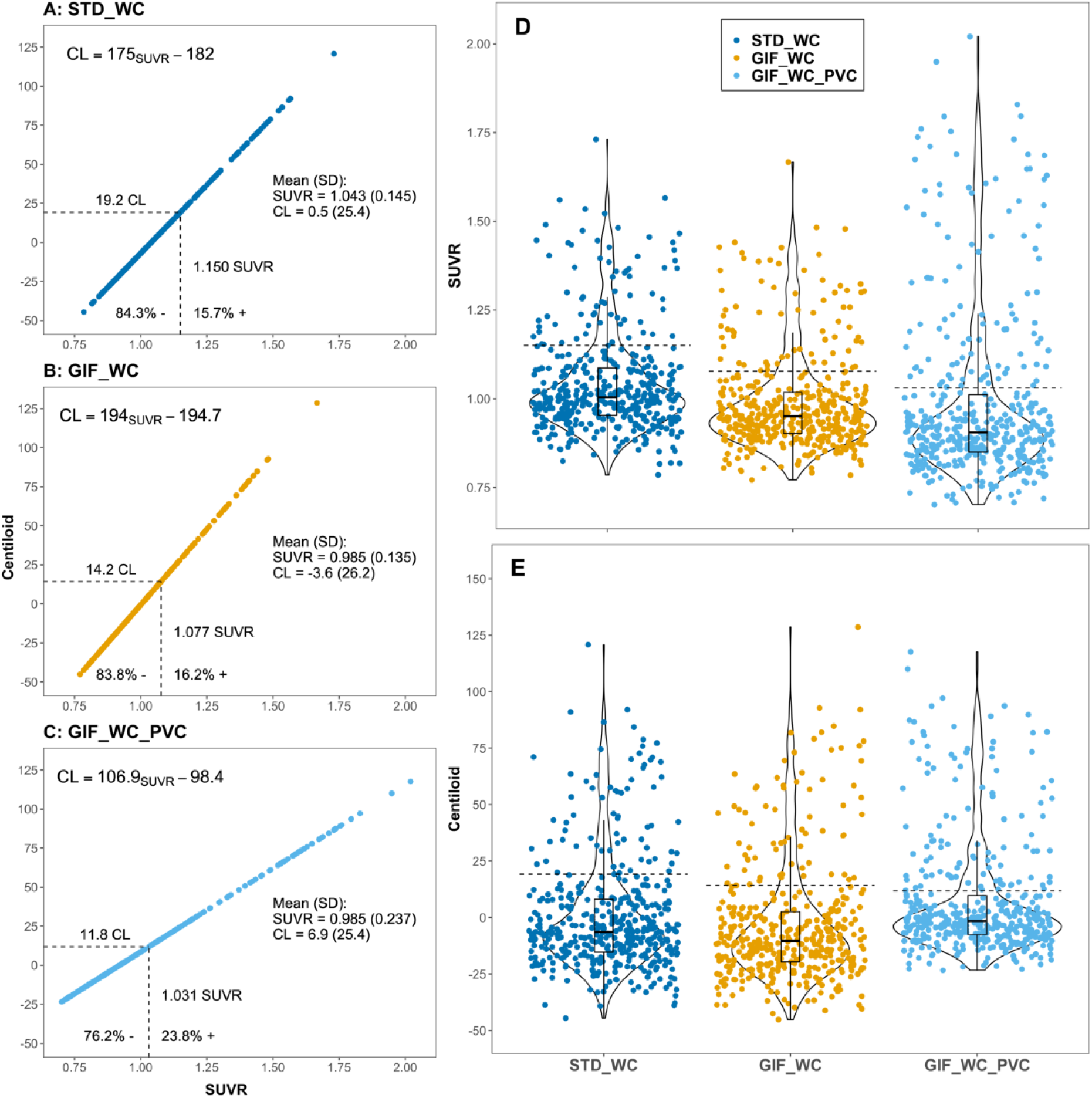
Centiloid conversion of florbetapir PET/MR data from Insight 46 processed using the whole cerebellum reference region. A-C: Plots showing the direct linear transformation from SUVR method to Centiloid values. Direct conversion equations shown for each method along with the percentage of Insight 46 participants (N = 432) classified as Aβ positive and negative. Dashed black lines represent cutpoint values derived using gaussian-mixture modelling in Insight 46. The distribution of SUVR (D) and Centiloid (E) for are shown for each processing method. Abbreviations: STD_WC = Standard Centiloid pipeline; GIF = Geodesic Information Flows pipeline; WC = whole cerebellum reference; PVC = partial volume corrected; SD = standard deviation; SUVR = standard uptake value ratio; CL = Centiloid units.

#### Eroded white matter reference region

The Aβ positivity rates were 18.3% and 18.1% for FBP_GIF_WM_SUVR_ and FBP_GIF_WM_PVC_SUVR_ pipelines, respectively. When the conversion procedure was applied to the FBP_GIF_WM_SUVR_ Insight 46 data, the SUVR cutpoint of 0.610 corresponded to -23.0 CL and the mean (SD) Centiloid value was -48.3 (39.5) (see Figure 5A). For FBP_GIF_WM_PVC_SUVR_, the SUVR cutpoint of 0.671 equated to +26.7 CL, with a mean (SD) Centiloid value of +10.5 (30.0) (see Figure 5C). Post-hoc analyses were performed to investigate these unexpected results.

**Figure 5.**
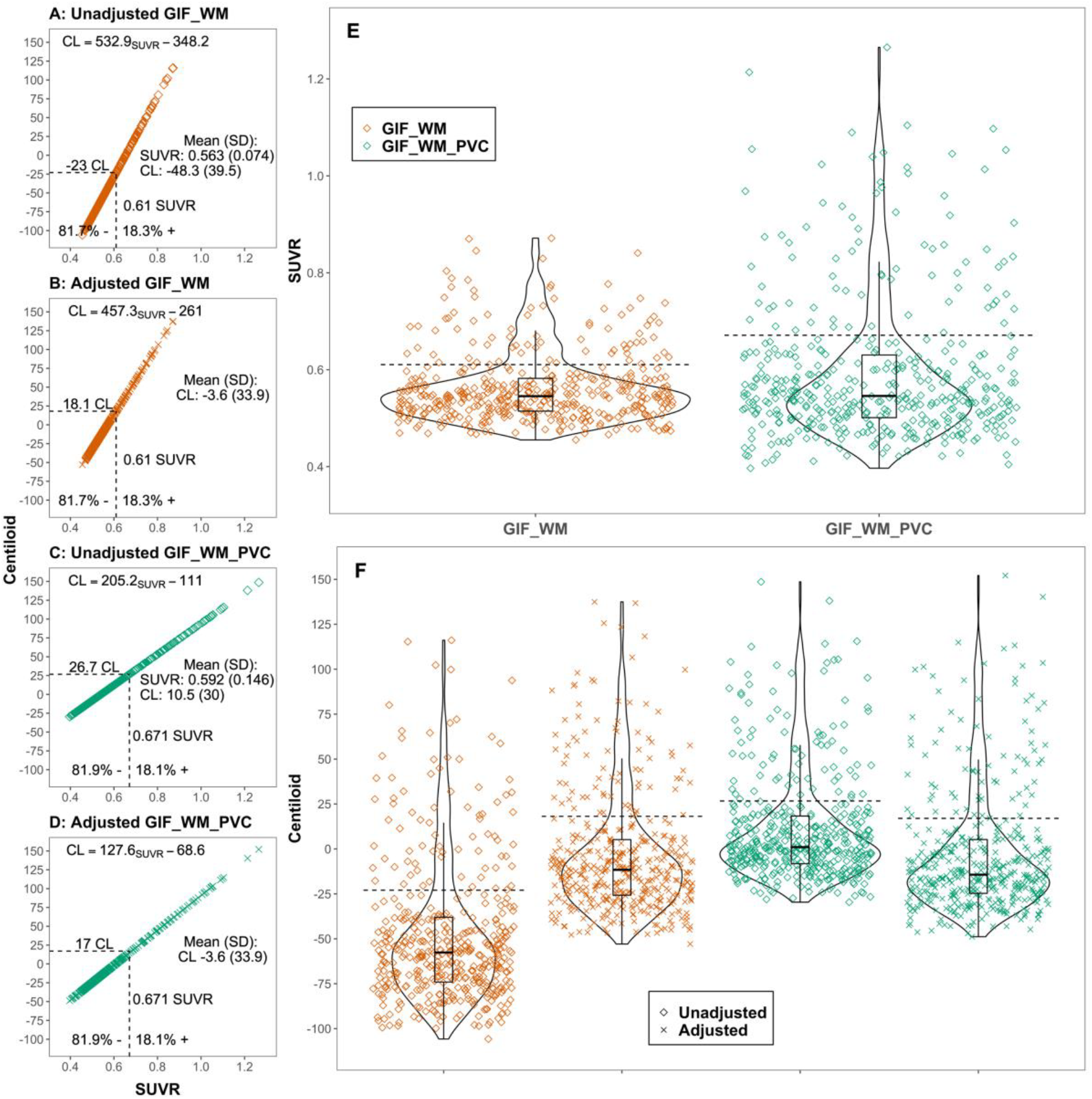
The Centiloid conversion of florbetapir PET/MR SUVRs from Insight 46 processed with an eroded white matter reference region, with unadjusted and adjusted Centiloid values. Dashed black lines represent cutpoint values derived using gaussian-mixture modelling in Insight 46. Abbreviations: STD_WC = Standard Centiloid pipeline; GIF = Geodesic Information Flows pipeline; WM = eroded white matter reference; PVC = partial volume corrected; SD = standard deviation; SUVR = standard uptake value ratio; CL = Centiloid units.

#### Adjustment for white matter SUVRs

Further exploration of the data indicated that there was a differential relationship between WM and WC uptake in Insight 46 compared to the Florbetapir Calibration dataset. The regression line between GIF_WM_SUVR_ (y) and STD_WC_SUVR_ (x) had a smaller slope and higher intercept in the Florbetapir Calibration (ŷ = 0.291x + 0.360, R^2^ = 0.67) compared to the Insight 46 dataset (ŷ = 0.388x + 0.159, R^2^ = 0.58). This differential relationship was also evident when comparing GIF_WM_SUVR_ (y) against GIF_WC_SUVR_ (x); see Figure 6. These values have identical target regions (defined by the GIF parcellation), with only the reference region changing. The red dashed lines on Figure 6A and B show the value of ^calc^GIF_WM_SUVR_ when GIF_WC_SUVR_ = 1 and demonstrates why the linear equation calculated in the Florbetapir Calibration dataset gives higher estimates of ^calc^GIF_WM_SUVR_ than is appropriate for the Insight 46 dataset. The reverse transformation from GIF_WM_SUVR_ to ^calc^GIF_WC_SUVR_ therefore leads to underestimated Centiloid values when converting GIF_WM_SUVR_ from Insight 46.

**Figure 6.**
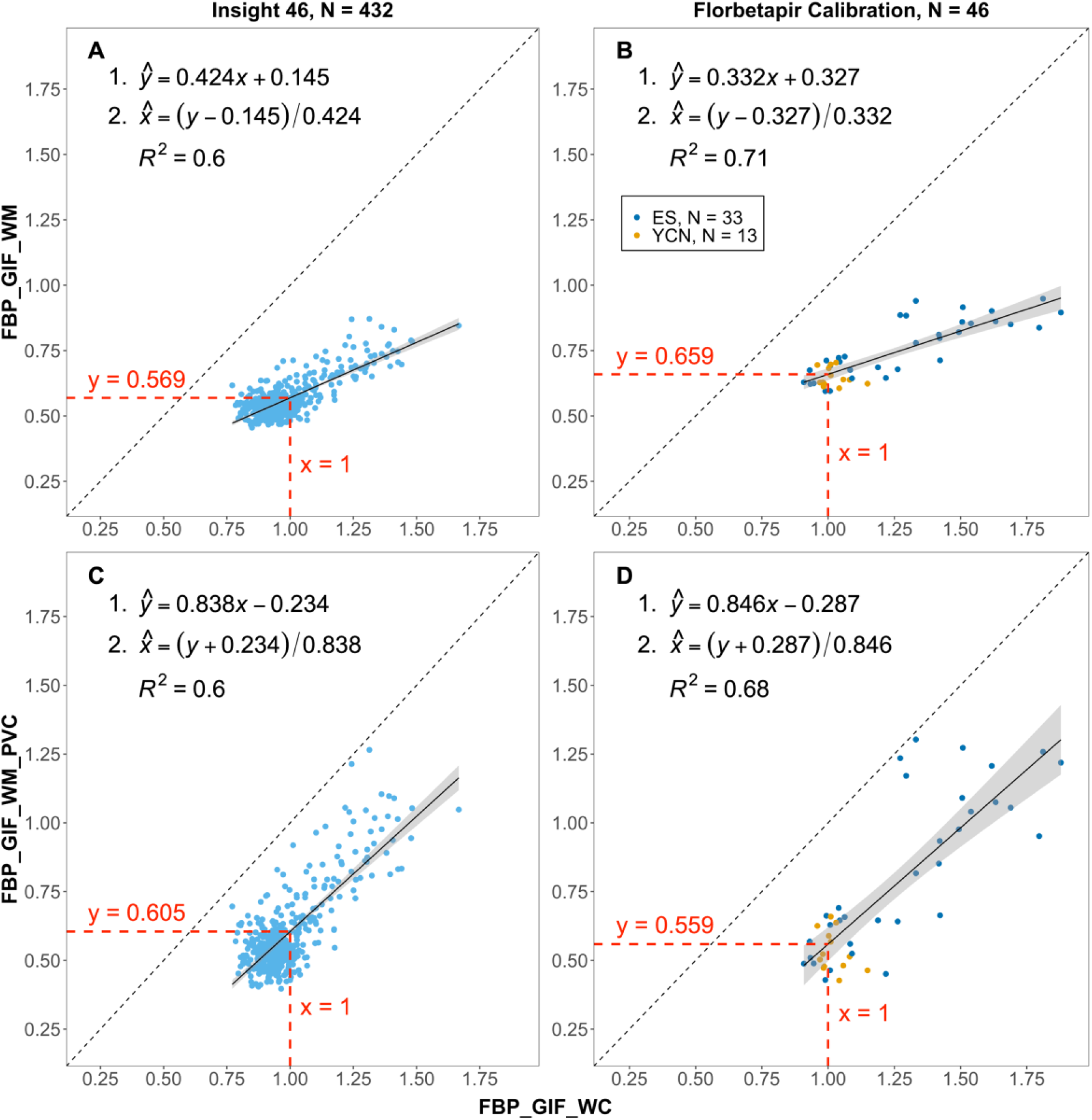
The differential relationship between GIF_WC and GIF_WM in with florbetapir data from the Insight 46 and Florbetapir Calibration datasets, leading to inappropriate scaling effects. Equation 2 on panel A and C was used to adjust the transforms for FBP_GIF_WM and FBP_GIF_WM_PVC, respectively. Red dashed lines represent the SUVR of y when GIF_WC = 1 for each relationship. Abbreviations: SUVR = Standardised uptake value ratio; FBP = florbetapir; GIF = Geodesic information flows pipeline; WC = Whole cerebellum reference; WM = Eroded white matter reference; PVC = Partial volume corrected; ES = Elder subjects; YCN = Young cognitively normal.

After conducting this additional analysis, we implemented a dataset-specific adjustment to convert WM normalised SUVRs from Insight 46 to Centiloids. As the Centiloid transformations for GIF_WC_SUVR_ appear to be generalisable to Insight 46, we added an initial step to convert GIF_WM_SUVR_ and GIF_WM_PVC_SUVR_ to ^calc^GIF_WC_SUVR_ (see Figure 1B).

The equations used for this step are equation 2 in Figures 6A and 6C, where 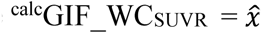. Following this adjustment, the FBP_GIF_WC_SUVR_ to Centiloid equation was applied to adjusted values (CL = ^calc^FBP_GIF_WC_SUVR_ × 194.0 – 194.7). The adjusted direct conversion equations and Centiloid cutpoints for FBP_GIF_WM_SUVR_ and FBP_GIF_WM_PVC_SUVR_ are presented in Figure 5B and 5D, respectively.

#### Supplementary analysis in ADNI dataset

In the ADNI dataset, the GMM derived cutpoint for FBP_GIF_WC_SUVR_ was 1.123 which scaled to 23 CL. For FBP_GIF_WM_SUVR_, the SUVR cutpoint was 0.691 which equated to 20.3 CL using the unadjusted centiloid transformation and 55.3 CL when instead using the adjusted equation. See section 4 of the supplementary materials for a direct comparison of the ratio of uptake in WM and WC reference regions between datasets.

## DISCUSSION

In this study, we found that florbetapir data acquired on a combined PET/MR scanner – in particular SUVR using a whole cerebellum reference - can successfully be transformed to the Centiloid scale. Following calibration of four non-standard SUVR methods to Centiloids, including the use of an eroded white matter reference region and PVC, we applied the scaling to florbetapir PET/MR data from a large cohort of ∼70-year-old individuals. We found that for SUVRs using a whole cerebellum reference region, Aβ positivity cutpoints fell within a range of 11.8 to 19.2 Centiloids. These gaussian-mixture modelling derived Centiloid cutpoint values are consistent with those reported by other studies using different methodologies [26–29]. However, the same conversion process when applied to eroded white matter reference SUVRs resulted in unexpected cutpoint values of -23.0 Centiloids without and +26.7 Centiloids with PVC. Having shown that these results appeared to be due to a differential relationship between white matter and whole cerebellum uptake in the Florbetapir Calibration dataset compared to the Insight 46 dataset, we introduced an adjustment step based on the relationship between uptake in reference regions in Insight 46 before scaling to Centiloids. This resulted in much more plausible cutpoint values of 18.1 without and 17.0 Centiloids with PVC.

The Centiloid method compresses or stretches results into a similar range, and it is important to report information regarding the reliability of the conversion and precision of each non-standard approach [7]. In the current study, we applied the Centiloid conversion to SUVRs with an eroded white matter reference region and with PVC applied. These extra degrees of separation from the Standard Centiloid processing approach could reduce the reliability of transformations. In the Standard PiB dataset, SUVRs from all GIF pipelines were strongly associated with the Standard Centiloid processing (all R^2^ > 0.90), indicating a reliable conversion between methods. The linear association between values from GIF whole cerebellum and Standard Centiloid pipelines was particularly strong (R^2^ = 0.99), with a slight underestimation for the GIF pipeline compared to the Standard Centiloid processing approach (Figure 2A). This underestimation could be due to greater inclusion of white matter uptake in the MNI space Standard Centiloid target region compared to the subject-specific native space cortical GIF target region. In the Florbetapir Calibration dataset, the conversion across both radiotracer and processing method were highly reliable for whole cerebellum regions (R^2^ > 0.87, Figures 3A and 3B) and lower, but still above the Centiloid threshold for white matter reference (R^2^ = 0.75, Figures 3C and 3D). The reliability of conversion was unaffected by application of PVC. In young controls presumed to have no Aβ accumulation, the relative variance between non-standard Centiloids compared to Standard PiB Centiloids reflects both the relative dynamic range and precision of the non-standard method [7]. Navitsky et al. [9] reported that florbetapir SUVRs had a dynamic range of about half that of PiB when using the Standard Centiloid processing, resulting in a doubling of the variance ratio between florbetapir and PiB SUVRs when scaling to Centiloids. Here, the dynamic range of florbetapir GIF whole cerebellum SUVRs was also around half of Standard PiB SUVRs (slope = 0.482 in Figure 3A). We found that a white matter reference reduced the dynamic range of florbetapir further (slope = 0.175 in Figure 3C) and PVC increased it for both whole cerebellum (slope = 0.874 in Figure 3B) and white matter referenced values (slope = 0.455 in Figure 3D). Relative variance was higher in florbetapir compared to PiB data and with white matter compared to whole cerebellum reference values (Table 2). GIF whole cerebellum with PVC had the lowest relative variance in florbetapir data, due to the increase in dynamic range without a meaningful increase in variability. The effect of different processing choices on variability were different for florbetapir and PiB SUVRs, as might be expected due to the increased off-target binding in white matter for florbetapir [6]. SUVR variability (CoV) was highest when using a white matter reference with PVC, although the increased range of values reduced the relative variance in Centiloids compared to non-PVC values. For consistency, the PVC parameters in the pipeline were kept consistent between datasets using the parameters optimised for the PET/MR florbetapir dataset. However it is important to note that the Florbetapir Calibration dataset is likely under corrected for partial volume effects. One reason is the lower spatial resolution of the older scanners used in the Florbetapir Calibration compared to the PET/MR dataset. Furthermore, the PiB scans will have slightly poorer resolution compared to florbetapir due to the higher energy of positrons in Carbon-11.

While Centiloid values can provide a more consistent measure of Aβ burden than SUVR, it is important to appreciate how scaled values are affected by the approach and dataset used [30]. In the current study, there was a lack of generalisability between datasets when converting SUVRs with an eroded white matter reference region, which we corrected for with a dataset specific linear adjustment. This adjustment does not change Aβ positivity rates but brings white matter values into the same range as whole cerebellum values before scaling to Centiloids. We hypothesise two potential sources of variation that could be contributing to the differences between datasets: (1) the method of image acquisition or reconstruction and (2) the biological characteristics of the cohorts. Neither of these are accounted for in the Centiloid approach, where equations are calculated on a calibration dataset and applied to an independent dataset. The Insight 46 dataset was acquired using a single PET/MR scanner with a large axial FoV (25.8 cm) compared to the scanners used for the Florbetapir Calibration and ADNI datasets, which were typically ∼16 cm [31]. Shorter bore scanners used in Centiloid and ADNI datasets are likely to have increased noise in peripheral brain structures, like the cerebellum, positioned near the edge of the axial FOV. Some studies have reported lower levels of longitudinal intra-individual variability with an eroded white matter reference, a relatively central structure compared to the cerebellum [4,16,19–22]. Therefore, it is possible that the ratio of cerebellar to white matter noise contributes to the differential relationship between SUVRs from datasets. However, the generalisability of the whole cerebellum results leads us to believe differences in white matter signal, rather than cerebellar noise, are likely playing a more substantial role in the bias. It is possible that differences in attenuation correction between PET/MR and PET/CT datasets could lead to some differences, although as attenuation correction differences have mainly been found in the cerebellum, we do not expect this to be causing the systematic bias observed with white matter reference values. Regarding the biological characteristics of the cohorts, age and disease status can affect radiotracer dynamics through changes in cerebral blood flow, which differentially affect cerebellum and white matter regions [17,32]. There are substantial sample differences between the Florbetapir Calibration dataset (a wide range of age and disease status from control to AD) and Insight 46 (community ageing cohort in tight age range 68-72) that could also contribute to the differential relationship between white matter and cerebellum uptake [33]. We aimed to address some of these differences with the ADNI (PET/CT) dataset of controls matched in age to Insight 46. We found the equations from the Florbetapir Calibration dataset to result in more appropriate white matter referenced Centiloids (cutpoint = 20.3 CL) compared to the values calculated using the adjustment derived from Insight 46 data (cutpoint = 55.3 CL). However, the unadjusted white matter mean of -3.1 CL is also lower than that of whole cerebellum referenced values in the ADNI dataset (FBP_GIF_WC mean = 12.9 CL), suggesting there is also some underlying difference between Florbetapir Calibration and ADNI. ADNI has stricter recruitment criteria against white matter disease compared to the Insight 46 community sample, which could result differences in florbetapir white matter uptake [34,35]. Several studies have attributed high Aβ tracer binding in white matter to an affinity for myelin basic protein, and have explored their use in multiple sclerosis [35–41]. This myelin involvement is a potential confounder of white matter as a reference region and could lead to differences in SUVR dependent on sex, age and white matter disease [40].

A limitation of this paper is that we are unable to identify the exact source of variation between datasets which leads to the lack of generalisable white matter referenced SUVRs. Ideally sources of variation from acquisition and sample could be characterised with further paired (PiB and florbetapir) calibration datasets controlling for all other factors; however, it is logistically difficult (time, cost, radiation regulation, radiotracer availability) to collect these data. We found a balance between logistics and generalisability by adjusting GIF white matter reference SUVR values to GIF whole cerebellum values within the Insight 46 dataset, then linking these values to the Centiloid scale using the generalisable whole cerebellum SUVR equations from the independent calibration dataset.

Future work will explore the Centiloid implementation with longitudinal follow-up data in this PET/MR dataset. The relationship between white matter and cerebellum florbetapir uptake will be examined further, which will be important for the standardisation of SUVR results using a white matter reference region.

## CONCLUSION

Here we explore the implementation of the Centiloid scale in a large florbetapir PET/MR dataset. We demonstrate that the conversion of whole cerebellum reference SUVRs, with and without PVC, can generalise to PET/MR datasets. We highlight the need for careful consideration of underlying differences between datasets that can affect values even after conversion to Centiloids, especially when using white matter reference regions and methodologies further removed from the Standard Centiloid approach. We show that a linear adjustment can facilitate conversion of values should differences between datasets arise.

## Data Availability

All data and scripts in the present study are available upon reasonable request to the authors or from publicly available datasets.

http://www.gaain.org/centiloid-project

http://adni.loni.usc.edu/

## REFERENCES

1. Jack CR, Knopman DS, Jagust WJ, Shaw LM, Aisen PS, Weiner MW, et al. Hypothetical model of dynamic biomarkers of the Alzheimer’s pathological cascade. Lancet Neurol. 2010;9:119–28.

2. Mintun MA, Lo AC, Duggan Evans C, Wessels AM, Ardayfio PA, Andersen SW, et al. Donanemab in Early Alzheimer’s Disease. N Engl J Med. 2021;NEJMoa2100708.

3. Lopresti BJ, Klunk WE, Mathis CA, Hoge JA, Ziolko SK, Lu X, et al. Simplified Quantification of Pittsburgh Compound B Amyloid Imaging PET Studies: A Comparative Analysis. J Nucl Med. Society of Nuclear Medicine; 2005;46:1959–72.

4. Chiao P, Bedell BJ, Avants B, Zijdenbos AP, Grand’Maison M, O’Neill P, et al. Impact of Reference and Target Region Selection on Amyloid PET SUV Ratios in the Phase 1b PRIME Study of Aducanumab. J Nucl Med. Society of Nuclear Medicine; 2019;60:100–6.

5. Landau SM, Thomas BA, Thurfjell L, Schmidt M, Margolin R, Mintun M, et al. Amyloid PET imaging in Alzheimer’s disease: a comparison of three radiotracers. Eur J Nucl Med Mol Imaging. 2014;41:1398–407.

6. Landau SM, Breault C, Joshi AD, Pontecorvo M, Mathis CA, Jagust WJ, et al. Amyloid-β Imaging with Pittsburgh Compound B and Florbetapir: Comparing Radiotracers and Quantification Methods. J Nucl Med. Society of Nuclear Medicine; 2013;54:70–7.

7. Klunk WE, Koeppe RA, Price JC, Benzinger TL, Devous MD, Jagust WJ, et al. The Centiloid Project: Standardizing quantitative amyloid plaque estimation by PET. Alzheimers Dement. 2015;11:1–15.e4.

8. Battle MR, Pillay LC, Lowe VJ, Knopman D, Kemp B, Rowe CC, et al. Centiloid scaling for quantification of brain amyloid with [18F]flutemetamol using multiple processing methods. EJNMMI Res. 2018;8:107.

9. Navitsky M, Joshi AD, Kennedy I, Klunk WE, Rowe CC, Wong DF, et al. Standardization of amyloid quantitation with florbetapir standardized uptake value ratios to the Centiloid scale. Alzheimers Dement. 2018;14:1565–71.

10. Rowe CC, Jones G, Doré V, Pejoska S, Margison L, Mulligan RS, et al. Standardized Expression of 18F-NAV4694 and 11C-PiB β-Amyloid PET Results with the Centiloid Scale. J Nucl Med. Society of Nuclear Medicine; 2016;57:1233–7.

11. Rowe CC, Doré V, Jones G, Baxendale D, Mulligan RS, Bullich S, et al. 18F-Florbetaben PET beta-amyloid binding expressed in Centiloids. Eur J Nucl Med Mol Imaging. 2017;44:2053–9.

12. Lane CA, Parker TD, Cash DM, Macpherson K, Donnachie E, Murray-Smith H, et al. Study protocol: Insight 46 – a neuroscience sub-study of the MRC National Survey of Health and Development. BMC Neurol. 2017;17:75.

13. Burgos N, Cardoso MJ, Thielemans K, Modat M, Pedemonte S, Dickson J, et al. Attenuation Correction Synthesis for Hybrid PET-MR Scanners: Application to Brain Studies. IEEE Trans Med Imaging. 2014;33:2332–41.

14. Cardoso MJ, Modat M, Wolz R, Melbourne A, Cash D, Rueckert D, et al. Geodesic Information Flows: Spatially-Variant Graphs and Their Application to Segmentation and Fusion. IEEE Trans Med Imaging. 2015;34:1976–88.

15. Modat M, Cash DM, Daga P, Winston GP, Duncan JS, Ourselin S. A symmetric block-matching framework for global registration. Med Imaging 2014 Image Process [Internet]. International Society for Optics and Photonics; 2014 [cited 2020 Nov 3]. p. 90341D. Available from: https://www.spiedigitallibrary.org/conference-proceedings-of-spie/9034/90341D/A-symmetric-block-matching-framework-for-global-registration/10.1117/12.2043652.short

16. Kameyama M, Ishibash K, Wagatsuma K, Toyohara J, Ishii K. A pitfall of white matter reference regions used in [18F] florbetapir PET: a consideration of kinetics. Ann Nucl Med. 2019;33:848–54.

17. Ottoy J, Verhaeghe J, Niemantsverdriet E, Wyffels L, Somers C, Roeck ED, et al. Validation of the Semiquantitative Static SUVR Method for 18F-AV45 PET by Pharmacokinetic Modeling with an Arterial Input Function. J Nucl Med. Society of Nuclear Medicine; 2017;58:1483–9.

18. Thal DR, Rüb U, Orantes M, Braak H. Phases of Aβ-deposition in the human brain and its relevance for the development of AD. Neurology. AAN Enterprises; 2002;58:1791–800.

19. Brendel M, Högenauer M, Delker A, Sauerbeck J, Bartenstein P, Seibyl J, et al. Improved longitudinal [18F]-AV45 amyloid PET by white matter reference and VOI-based partial volume effect correction. NeuroImage. 2015;108:450–9.

20. Chen K, Roontiva A, Thiyyagura P, Lee W, Liu X, Ayutyanont N, et al. Improved Power for Characterizing Longitudinal Amyloid-β PET Changes and Evaluating Amyloid-Modifying Treatments with a Cerebral White Matter Reference Region. J Nucl Med. Society of Nuclear Medicine; 2015;56:560–6.

21. Fleisher AS, Joshi AD, Sundell KL, Chen Y-F, Kollack-Walker S, Lu M, et al. Use of white matter reference regions for detection of change in florbetapir positron emission tomography from completed phase 3 solanezumab trials. Alzheimers Dement. 2017;13:1117–24.

22. Landau SM, Fero A, Baker SL, Koeppe R, Mintun M, Chen K, et al. Measurement of Longitudinal β-Amyloid Change with 18F-Florbetapir PET and Standardized Uptake Value Ratios. J Nucl Med. Society of Nuclear Medicine; 2015;56:567–74.

23. Erlandsson K, Buvat I, Pretorius PH, Thomas BA, Hutton BF. A review of partial volume correction techniques for emission tomography and their applications in neurology, cardiology and oncology. Phys Med Biol. IOP Publishing; 2012;57:R119–59.

24. Thomas BA, Erlandsson K, Modat M, Thurfjell L, Vandenberghe R, Ourselin S, et al. The importance of appropriate partial volume correction for PET quantification in Alzheimer’s disease. Eur J Nucl Med Mol Imaging. 2011;38:1104–19.

25. Hutton BF, Thomas BA, Erlandsson K, Bousse A, Reilhac-Laborde A, Kazantsev D, et al. What approach to brain partial volume correction is best for PET/MRI? Nucl Instrum Methods Phys Res Sect Accel Spectrometers Detect Assoc Equip. 2013;702:29–33.

26. Farrell ME, Jiang S, Schultz AP, Properzi MJ, Price JC, Becker JA, et al. Defining the Lowest Threshold for Amyloid-PET to Predict Future Cognitive Decline and Amyloid Accumulation. Neurology. Wolters Kluwer Health, Inc. on behalf of the American Academy of Neurology; 2021;96:e619–31.

27. Jack CR, Wiste HJ, Weigand SD, Therneau TM, Lowe VJ, Knopman DS, et al. Defining imaging biomarker cut points for brain aging and Alzheimer’s disease. Alzheimers Dement. 2017;13:205–16.

28. La Joie R, Ayakta N, Seeley WW, Borys E, Boxer AL, DeCarli C, et al. Multisite study of the relationships between antemortem [11C]PIB-PET Centiloid values and postmortem measures of Alzheimer’s disease neuropathology. Alzheimers Dement. 2019;15:205–16.

29. Salvadó G, Molinuevo JL, Brugulat-Serrat A, Falcon C, Grau-Rivera O, Suárez-Calvet M, et al. Centiloid cut-off values for optimal agreement between PET and CSF core AD biomarkers. Alzheimers Res Ther. 2019;11:27.

30. Su Y, Flores S, Hornbeck RC, Speidel B, Vlassenko AG, Gordon BA, et al. Utilizing the Centiloid scale in cross-sectional and longitudinal PiB PET studies. NeuroImage Clin. 2018;19:406–16.

31. Delso G, Fürst S, Jakoby B, Ladebeck R, Ganter C, Nekolla SG, et al. Performance Measurements of the Siemens mMR Integrated Whole-Body PET/MR Scanner. J Nucl Med. Society of Nuclear Medicine; 2011;52:1914–22.

32. van Berckel BNM, Ossenkoppele R, Tolboom N, Yaqub M, Foster-Dingley JC, Windhorst AD, et al. Longitudinal Amyloid Imaging Using 11C-PiB: Methodologic Considerations. J Nucl Med. Society of Nuclear Medicine; 2013;54:1570–6.

33. Lowe VJ, Lundt ES, Senjem ML, Schwarz CG, Min H-K, Przybelski SA, et al. White Matter Reference Region in PET Studies of 11C-Pittsburgh Compound B Uptake: Effects of Age and Amyloid-β Deposition. J Nucl Med. Society of Nuclear Medicine; 2018;59:1583–9.

34. Brickman AM, Zahra A, Muraskin J, Steffener J, Holland CM, Habeck C, et al. Reduction in cerebral blood flow in areas appearing as white matter hyperintensities on magnetic resonance imaging. Psychiatry Res Neuroimaging. 2009;172:117–20.

35. Pietroboni AM, Carandini T, Colombi A, Mercurio M, Ghezzi L, Giulietti G, et al. Amyloid PET as a marker of normal-appearing white matter early damage in multiple sclerosis: correlation with CSF β-amyloid levels and brain volumes. Eur J Nucl Med Mol Imaging. 2019;46:280–7.

36. Auvity S, Tonietto M, Caillé F, Bodini B, Bottlaender M, Tournier N, et al. Repurposing radiotracers for myelin imaging: a study comparing 18F-florbetaben, 18F-florbetapir, 18F-flutemetamol,11C-MeDAS, and 11C-PiB. Eur J Nucl Med Mol Imaging. 2020;47:490–501.

37. Bodini B, Veronese M, García-Lorenzo D, Battaglini M, Poirion E, Chardain A, et al. Dynamic Imaging of Individual Remyelination Profiles in Multiple Sclerosis. Ann Neurol. 2016;79:726–38.

38. Carotenuto A, Giordano B, Dervenoulas G, Wilson H, Veronese M, Chappell Z, et al. [18F]Florbetapir PET/MR imaging to assess demyelination in multiple sclerosis. Eur J Nucl Med Mol Imaging. 2020;47:366–78.

39. de Paula Faria D, Copray S, Sijbesma JWA, Willemsen ATM, Buchpiguel CA, Dierckx RAJO, et al. PET imaging of focal demyelination and remyelination in a rat model of multiple sclerosis: comparison of [11C]MeDAS, [11C]CIC and [11C]PIB. Eur J Nucl Med Mol Imaging. 2014;41:995–1003.

40. Moscoso A, Whitman A, Baker SL, La Joie R, Pascoal TA, Rosa-Neto P, et al. Reduced [18 F] flortaucipir retention in white matter hyperintensities compared to normal-appearing white matter. ALZ; 2020.

41. Stankoff B, Freeman L, Aigrot M-S, Chardain A, Dollé F, Williams A, et al. Imaging central nervous system myelin by positron emission tomography in multiple sclerosis using [methyl-11C]-2-(4′-methylaminophenyl)-6-hydroxybenzothiazole. Ann Neurol. 2011;69:673–80.

