## Supplementary Materials for "Operationalising the Centiloid Scale for [^18^F]florbetapir PET Studies on PET/MR"

### Operationalising the Centiloid Scale for [18F]florbetapir PET Studies on PET/MR: Supplementary Materials

#### Section 1. Sample Attrition

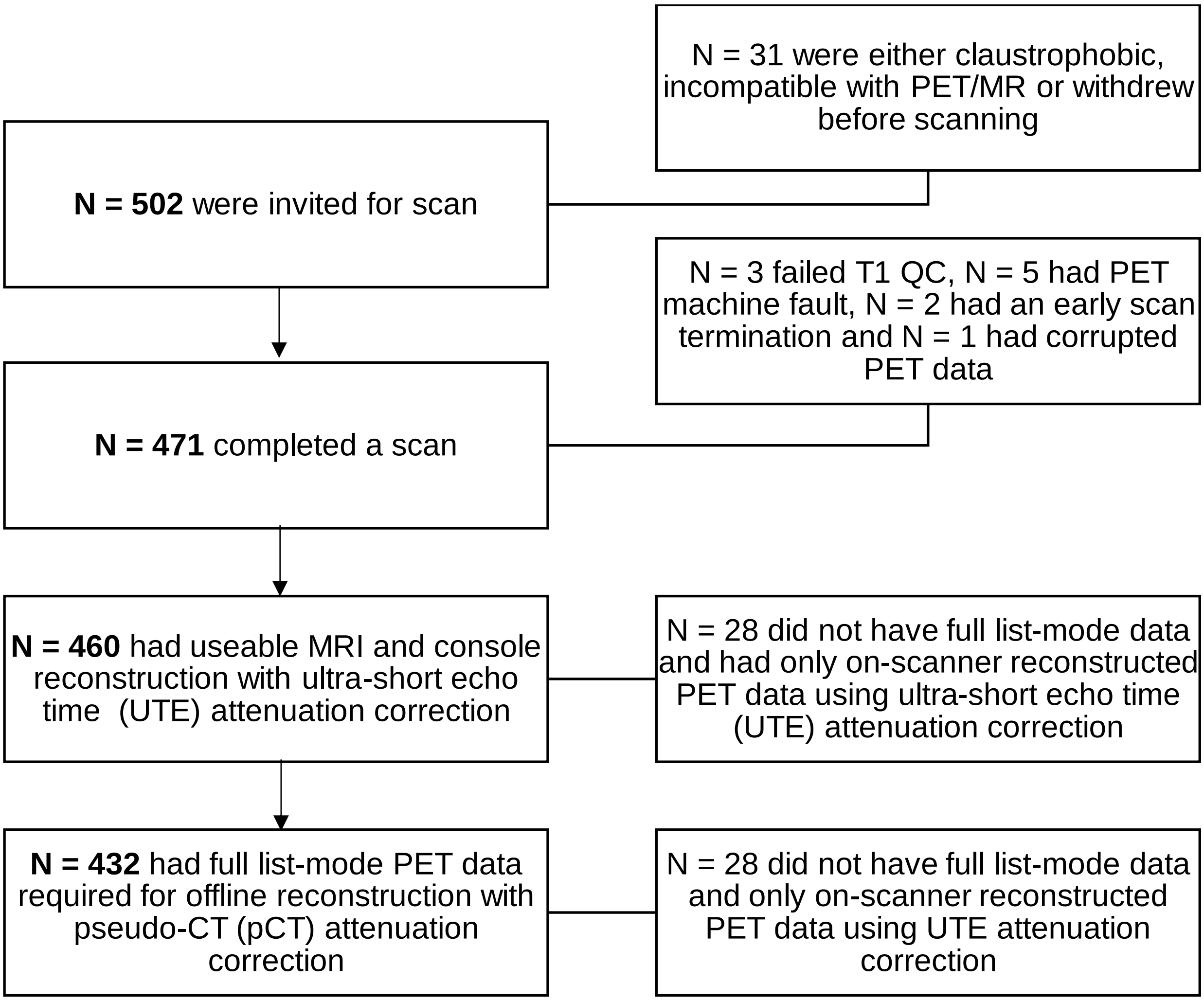


#### Section 2. pCT vs UTE reconstruction for PET/MR

We used pCT reconstruction in this study, SUVR results are highly correlated with results using ultrashort echo-time (UTE) MR attenuation correction, Figure 1.

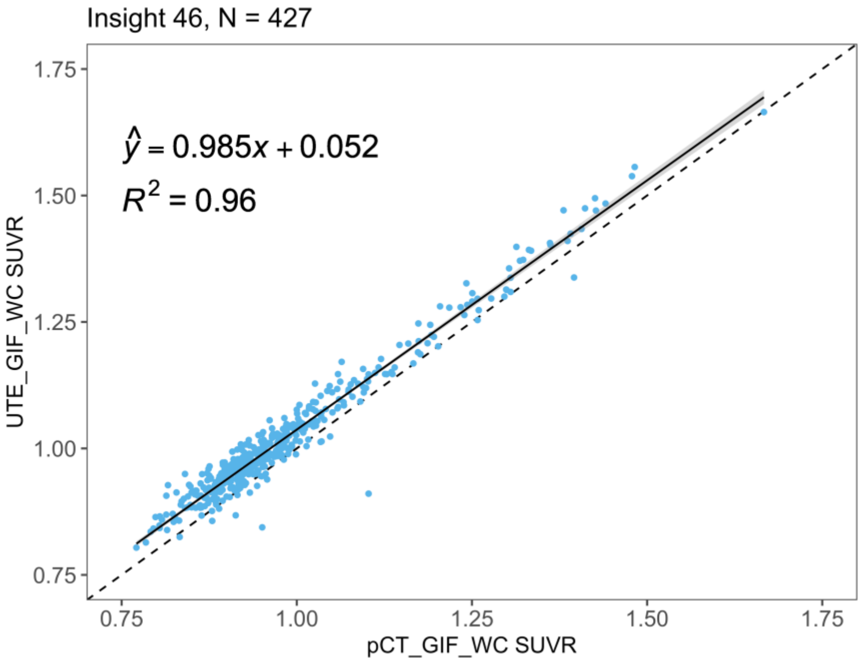


*Figure 1. Pseudo-CT (pCT) vs ultrashort echo-time (UTE) reconstructed SUVRs, processed using the geodesic information flows (GIF) pipeline with whole cerebellum (WC) reference, without partial volume correction.*

#### Section 3. Level 1 replication analysis and non-standard pipelines in the Standard PiB dataset

We downloaded the Standard PiB 50-70 min dataset and processed with SPM8 according to the Centiloid standard processing (PiB_STD_WC$_{SUVR}$) and compared to published values ($^{PUB}$PiB_STD_WC$_{SUVR}$) from Klunk et al. (2015). Replicate SUVRs should fall within 2% of published means for both the YC-0 (mean = 1.009, +/- 2% = 0.990 to 1.029) and AD-100 groups (mean = 2.076, +/- 2% = 2.056 to 2.096) groups. Our replicate pipeline ($^{REP}$PiB_STD_WC$_{SUVR}$) produced a YC-0 group mean (SD) = 1.014 (0.048) and AD-100 group mean (SD) = 2.087 (0.207), falling within the limits.

$^{REP}$PiB_STD_WC$_{SUVR}$ group means were then used as anchor points to scale our SUVRs from the Standard PiB dataset (Eq. 1), according to the level 1 replication analysis guidelines (Klunk et al., 2015).

Eq. 1.

$$^{REP}PiB\_STD\_WC_{CL} = 100 \times \frac{(^{REP}PiB\_STD\_WC_{SUVR} - 1.014)}{(2.087 - 1.014)}$$

The slope should be between 0.98 and 1.02, intercept between -2 and 2 CL and $R^2 > 0.98$. Figure 2 shows that the replicate values satisfy these criteria.

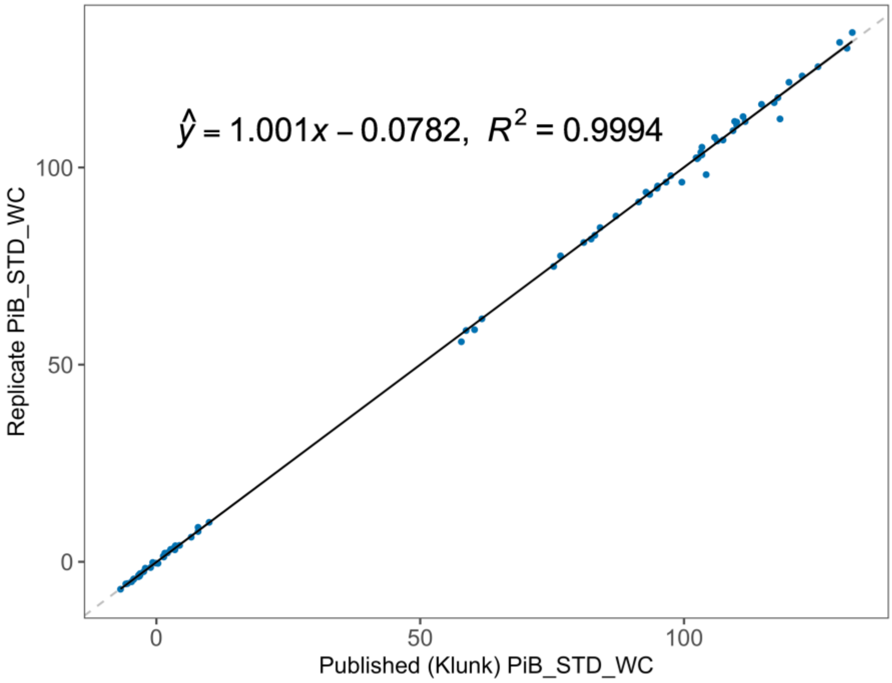


*Figure 2. Replicated Centiloid values (y) and published Centiloid values (x) using the standard whole cerebellum processing method (PiB_STD_WC) on the standard PiB dataset.*

Next, SUVRs from each GIF pipeline (y) were regressed against $^{REP}$PiB_STD_WC$_{SUVR}$, converted to $^{calc}$PiB_STD_WC$_{SUVR}$, then scaled using Sup Eq.1. The results of this are displayed in Figure 3.

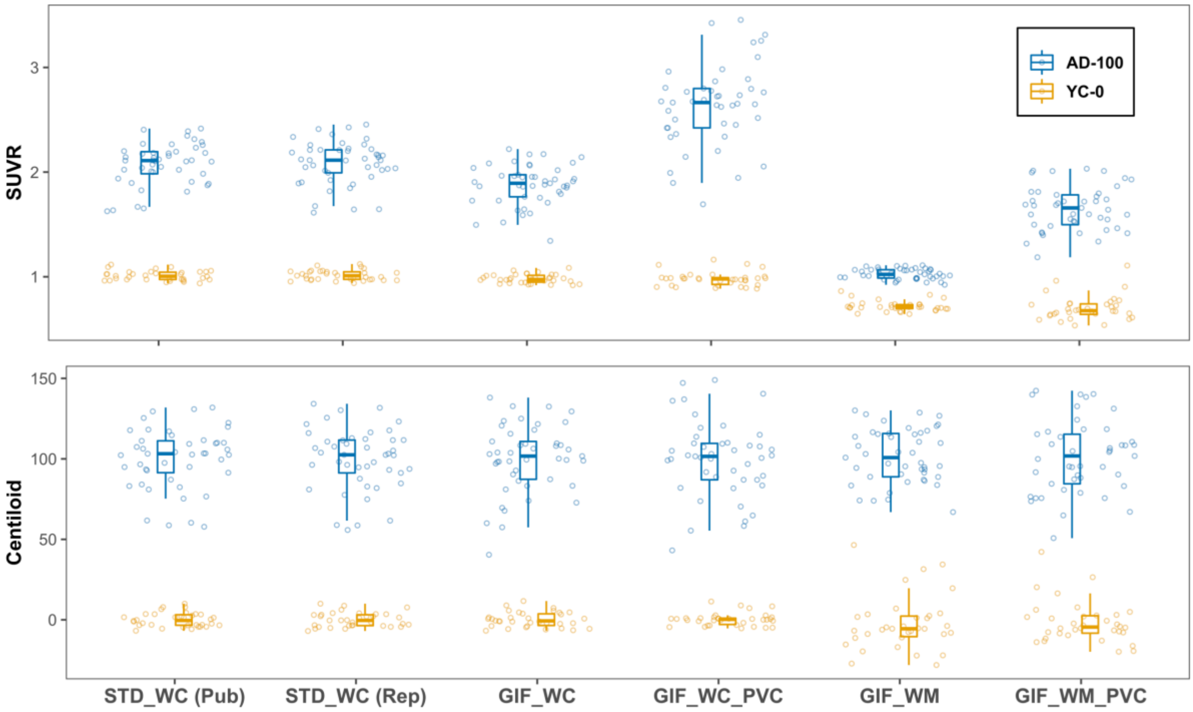


*Figure 3. SUVR and Centiloid values for all processing methods in the Standard PiB dataset. The transformation brings the methods into a comparable dynamic range.*

*Table 1. The variability of from all processing pipelines in the Standard PiB dataset, the coefficient of variation (CoV) for the young control (YC-0) group and effect size between groups.*

*STD_WC = standard Centiloid pipeline; GIF = geodesic information flows pipeline; WC = whole cerebellum reference; WM = eroded white matter reference; PVC = partial volume corrected; YC-0 = young control group; AD-100 = typical Alzheimer's disease group; PiB = Pittsburgh Compound B; SD = standard deviation; SUVR = standard uptake value ratio; CoV = Coefficient of Variance.*

| | | | PUBSTD_WC | GIF_WC | GIF_WC_PVC | GIF_WM | GIF_WM_PVC |
|---|---|---|---|---|---|---|---|
| **Standard PiB** | **YC-0 (N = 34)** | **Mean** | 1.01 | 0.98 | 0.98 | 0.72 | 0.70 |
| | | **SD** | 0.05 | 0.04 | 0.07 | 0.05 | 0.12 |
| | | **CoV** | 4.6 | 4.3 | 6.8 | 7.0 | 16.9 |
| | **AD-100 (N = 45)** | **Mean** | 2.08 | 1.88 | 2.64 | 1.03 | 1.65 |
| | | **SD** | 0.19 | 0.19 | 0.39 | 0.05 | 0.21 |
| | **Effect Size** | | 6.88 | 6.08 | 5.53 | 6.18 | 5.41 |

#### Section 4. Analysis of ADNI control FBP dataset

We carried out a supplementary analysis with the aim of replicating our finding of an inappropriate scaling effect for SUVRs normalised to eroded WM uptake in another independent dataset. We hypothesised that the differential relationship between WC uptake and WM uptake in the Florbetapir Calibration and Insight 46 datasets could arise from either 1) differences in the sample demographic, for example age or disease status, leading to differences in WM florbetapir uptake or 2) differences in acquisition, for example PET/MR vs PET or PET/CT, or some combination of the two factors.

To explore the generalisability of the Centiloid conversion on a sample that is more comparable to Insight 46 than the Florbetapir Calibration dataset we used, PET/CT data from ADNI controls of a similar age to Insight 46.

##### Methods

We downloaded florbetapir-PET (FBP) and a T1-weighted MRI scans for N = 93 ADNI control participants between the age of 68-72 years. Only participants with MRI acquired within one year of the PET were included, in order to minimise partial volume effects induced by atrophy mismatch between PET and MRI.

We did not use the fully preprocessed ADNI FBP images, as these are 50-70 mins post-injection, compared to 50-60 mins used in the Florbetapir Calibration and Insight 46 datasets.

ADNI images were downloaded in the ‘Co-registered Dynamic’ form and the first two frames were averaged (50-60 min), no other preprocessing steps were conducted. These ADNI scans were then processed using the GIF pipelines, with whole cerebellum (WC) and eroded white matter (WM) reference regions, both with and without partial volume correction (PVC).

Gaussian-mixture modelling (GMM) was performed on the ADNI dataset to calculate equivalent cutpoints to those already calculated in the Insight 46 dataset, see Figure 4.

**Results**

SUVR and Centiloid values for ADNI are presented in Table 2. Cutpoints and positivity rates defined using GMM on either Insight 46 or ADNI (Figure 4) datasets are compared.

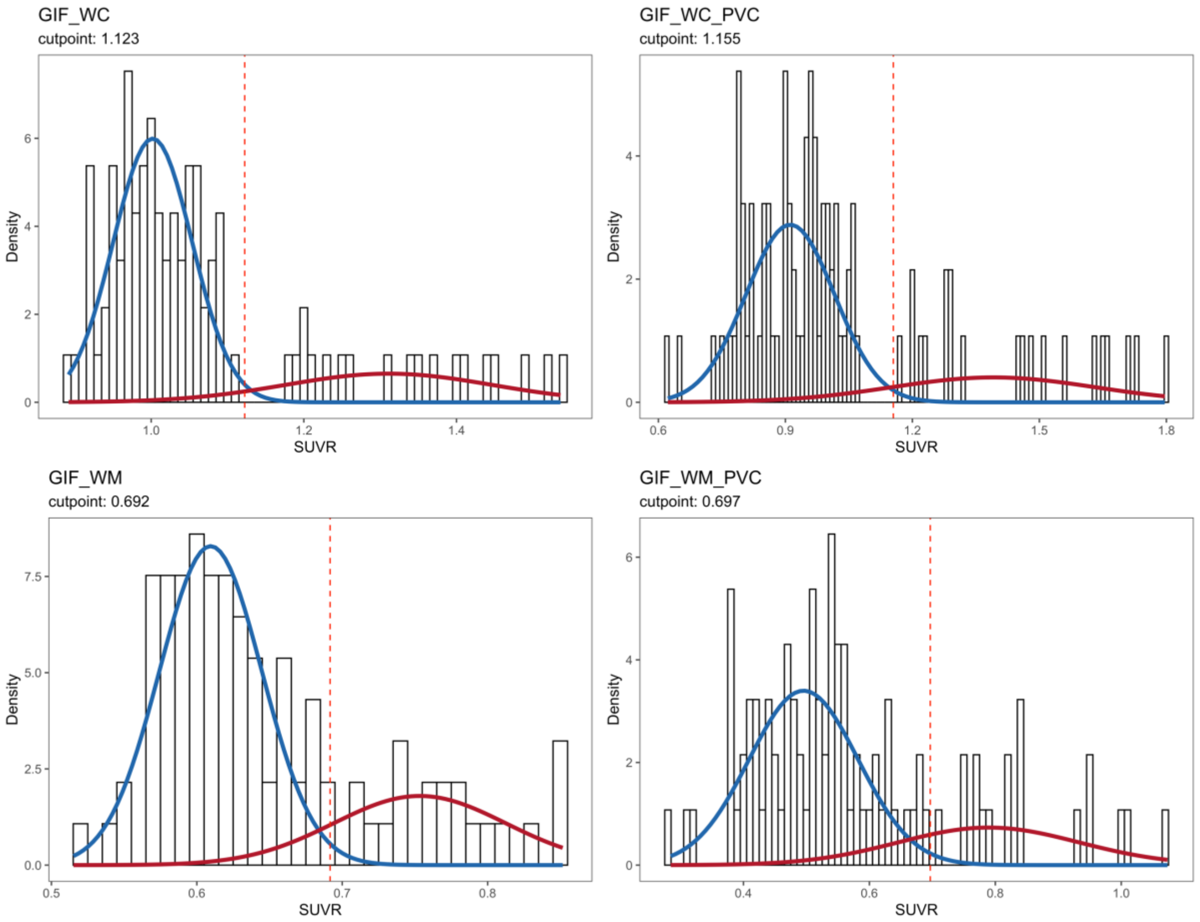


*Figure 4. GMM on the ADNI control dataset, N=93. The cutpoint was defined as 99th percentile of the negative distribution, the threshold was used in the Insight 46 dataset.*

*Table 2. SUVR and Centiloid values from the FBP dataset of N = 93 ADNI controls aged 60-72 years. Cutpoints derived from Gaussian-mixture modelling (two clusters, 99$^{th}$ percentile of lower distribution) on Insight 46 and the ADNI dataset are compared.*

| | | GIF_WC | GIF_WC_PVC | GIF_WM | GIF_WM Adjusted CL | GIF_WM_PVC | GIF_WM_PVC Adjusted CL |
|---|---|---|---|---|---|---|---|
| **Mean SUVR (SD)** | | 1.070 (0.151) | 1.028 (0.256) | 0.648 (0.077) | - | 0.573 (0.167) | - |
| **Mean Centiloid (SD)** | | 12.9 (29.3) | 11.6 (27.4) | -3.1 (41.1) | 35.2 (35.3) | 6.5 (34.3) | 4.5 (21.3) |
| **Cutpoint SUVR (CL)** | **Insight 46 Derived** | 1.077 (14.2) | 1.031 (11.8) | 0.610 (-23.0) | 0.610 (18.1) | 0.671 (26.7) | 0.671 (17.0) |
| | **ADNI Derived** | 1.123 (23.0) | 1.155 (25.1) | 0.691 (20.3) | 0.691 (55.3) | 0.697 (32.0) | 0.697 (20.3) |
| **Amyloid Status N+/N- (%+)** | **Insight 46 Derived** | 26/67 (28.0) | 28/65 (30.1) | 56/37 (60.2) | - | 21/72 (22.6) | - |
| | **ADNI Derived** | 19/74 (20.4) | 21/72 (22.6) | 20/73 (21.5) | - | 18/75 (19.4) | - |

WM to WC ratio

The WM to WC uptake ratio (GIF_WM_WC$_{SUVR}$) was then calculated for each individual using Eq. 2 below:

Eq. 2.

$$\frac{GIF\_WC_{SUVR}}{GIF\_WM_{SUVR}} = \frac{WM_{uptake}}{WC_{uptake}} = GIF\_WM\_WC_{SUVR}$$

Where 'GIF_WC$_{SUVR}$' is the GIF cortical composite target region uptake normalised by WC, and 'GIF_WC$_{SUVR}$' is GIF composite normalised by eroded WM.

Shapiro-Wilk normality tests performed on GIF_WM_WC$_{SUVR}$ in the three datasets showed the Florbetapir Calibration and ADNI datasets were non-normally distributed ($p$ = .029 and $p$ = .004, respectively). GIF_WM_WC$_{SUVR}$ in the Insight 46 dataset were normally distributed ($p$ = .830).

Non-parametric Wilcoxon rank-sum tests were used to compare each dataset to the Insight 46 dataset. The mean GIF_WM_WC$_{SUVR}$ was 5.8% lower both the ADNI (N = 93, mean = 1.654, SD = 0.135, $p < .001$) and Florbetapir Calibration datasets (N = 46, mean = 1.655, SD = 0.200, $p < .001$) relative to the Insight 46 dataset (N = 432, mean = 1.756, SD = 0.135).
